# An Ensemble Model for Acute Myeloid Leukemia Risk Stratification Recommendations by Combining Machine Learning with Clinical Guidelines

**DOI:** 10.1101/2024.01.08.24301018

**Authors:** Ming-Siang Chang, Xavier Cheng-Hong Tsai, Wen-Chien Chou, Hwei-Fang Tien, Hsin-An Hou, Chien-Yu Chen

## Abstract

Acute Myeloid Leukemia (AML) is a complex disease requiring accurate risk stratification for effective treatment planning. This study introduces an innovative ensemble machine learning model integrated with the European LeukemiaNet (ELN) 2022 recommendations to enhance AML risk stratification. The model demonstrated superior performance by utilizing a comprehensive dataset of 1,213 patients from National Taiwan University Hospital (NTUH) and an external cohort of 2,113 patients from UK-NCRI trials. On the external cohort, it improved a concordance index (c-index) from 0.61 to 0.64 and effectively distinguished three different risk levels with median hazard ratios ranging from 18% to 50% improved. Key insights were gained from the discovered significant features influencing risk prediction, including age, genetic mutations, and hematological parameters. Notably, the model identified specific cytogenetic and molecular alterations like *TP53, IDH2, SRSF2, STAG2, KIT, TET2*, and karyotype (-5, -7, -15, inv(16)), alongside age and platelet counts. Additionally, the study explored variations in the effectiveness of hematopoietic stem cell transplantation (HSCT) across different risk levels, offering new perspectives on treatment effects. In summary, this study develops an ensemble model based on the NTUH cohort to deliver improved performance in AML risk stratification, showcasing the potential of integrating machine learning techniques with medical guidelines to enhance patient care and personalized medicine.

## 1. Introduction

Acute Myeloid Leukemia (AML) is a rapidly progressing cancer of the blood and bone marrow, necessitating precise risk stratification for personalized treatment strategies.^1^ Traditional methods often relied on single or limited biomarkers for risk assessment. However, the complexity of AML shifted the focus towards integrating a variety of biomarkers for more nuanced risk evaluation.

The European LeukemiaNet (ELN) recommendations for AML utilize biomarkers like cytogenetics and gene mutations to categorize patients into different risk groups, ^2;3^ acknowledging the complex interactions between these biomarkers. For instance, the presence of the t(9;11)(p21.3;q23.3) chromosomal abnormality, which classifies a patient at an intermediate risk level, is prioritized over other rare, concurrent adverse gene mutations like ASXL1 in risk assessment. Besides, patients with *CEBPA* gene mutations are considered favorable in ELN but will result in poor prognosis if they carry *WT1* gene mutations simultaneously.^4^ Although previous studies have highlighted the benefits of accounting for the interactions between various biomarkers, they have not fully utilized the complexity of these relationships. Therefore, developing a more comprehensive model that can leverage multiple biomarkers simultaneously is necessary.

In medical machine learning (ML) models for risk stratification, clinician-initiated and non-clinician-initiated data play distinct roles. ^5^ Clinician-initiated data such as medical prescriptions, although helpful, might lead models to mimic existing clinical decisions and reduce the actual effect of risk stratification models, which aim to identify patients’ risk levels before clinical diagnosis. For instance, while the risk stratification model significantly outperforms the ELN model, it relies on data about whether to proceed with a transplant. ^6^ This data generation occurs after a doctor confirms risk, thus losing the original intent of the model to predict risk. Therefore, using non-clinician-initiated data is crucial for maintaining the integrity of risk stratification models. These data types, including patient-reported outcomes, vital signs monitored, and other health indicators, are gathered independently of clinical decisions. They provide a more unbiased basis for assessing patient risk. By incorporating this data, models can better fulfill their purpose of early risk identification rather than merely reflecting decisions already made by clinicians.

This study aims to enhance AML risk stratification at diagnosis by developing an ensemble machine learning model that combines predictions with the European LeukemiaNet (ELN) 2022 recom-mendations. ^2;3^ The model utilizes multiple non-clinician-initiated biomarkers, such as age, gender, hematologic data, karyotypes, and gene mutations, to accurately differentiate risk levels.

## 2. Methods

This study employed datasets from 1,213 patients from the National Taiwan University Hospital (NTUH) AML cohort and another from 2,113 patients from three UK-NCRI trials as an external cohort. ^7;8;9;10^ The NTUH cohort was filtered based on specific criteria and divided into training and validation sets to establish the proposed models. The models involved many well-known classification techniques. In addition, HyperOpt, ^11^ a Bayesian hyperparameter optimization, was used to select the optimal hyperparameters of each model. Finally, an ensemble (ML) model considers all classification techniques to predict risk. After integrating the classification techniques, we jointly consider the results by Ensemble (ML) and the results predicted by ELN 2022. Based on this, the study proposed clinical risk stratification recommendations (Ensemble (ML+ELN)).

### 2.1. Preprocessing the dataset for establishing models

The dataset for establishing models came from an acute myeloid leukemia cohort with 1,213 samples in NTUH, including age, gender, hematologic data, karyotypes, and gene mutations. After excluding those who did not get standard therapy and those who survived with follow-up time that did not surpass 36 months, the remaining 801 samples were separated into an 80% train set and 20% validation set for prediction and evaluation (Fig. 1). The exclusion is necessary as we can not confirm whether the short follow-up period leads to patients being labeled a poor risk. The labeling method of samples was defined by overall survival as follows (Table 1):

**Table 1.** The distribution of overall survival label for 801 samples.

| Overall survival label | Number (n=801) | Percentage (%) |
| --- | --- | --- |
| Adverse (< 12 months) | 254 | 31.71 |
| Intermediate | 282 | 35.21 |
| Favorable (> 60 months) | 265 | 33.08 |

**Figure 1.**
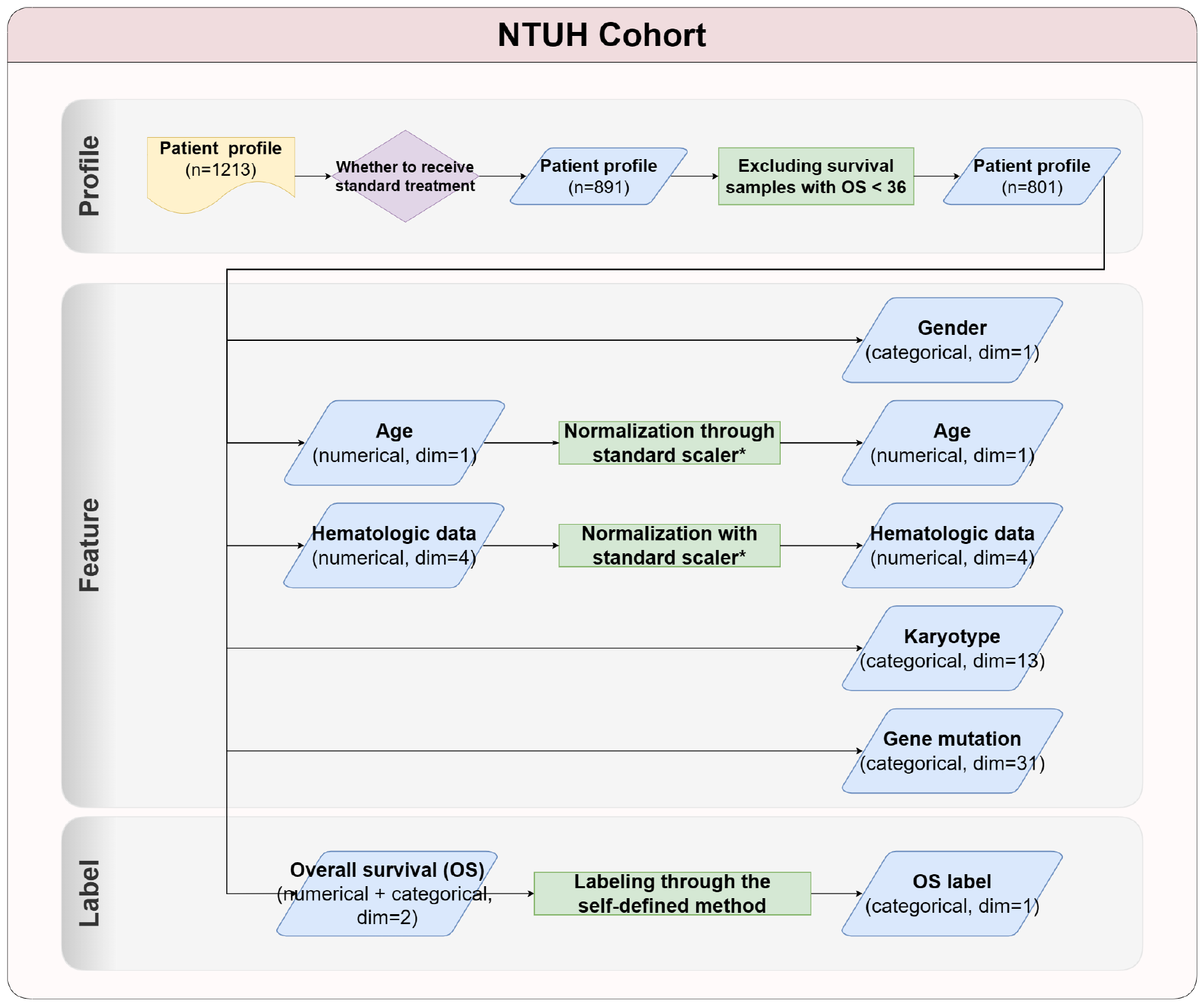
Feature selection and data preprocessing on the NTUH cohort. *Standard scaler is a normalization method in scikit-learn.^13^

- Less than 12 months was labeled as an adverse risk.
- Between 12 to 60 months was labeled as an intermediate risk.
- More than 60 months was labeled as a favorable risk.

The 13 types of karyotypes on the ELN recommendations for initial diagnosis were selected as features. ^2;3^ In addition, age, gender, four types of hematological data, and 31 types of gene mutations were also included. A previous study showed that the preprocessing techniques employed have no significant effect when none of the features have more than 10% missing values. ^12^ The standard normalization was applied to age and hematological data. ^13^ The data preprocessing and feature selection are shown in Fig. 1.

### 2.2. Models

The study employed various machine learning models with unique strengths and learning the relationships between features and labels. ^13^ Logistic Regression estimates the logistic model’s parameters to predict the likelihood of an event by making its log odds a linear combination of independent variables. ^14^ It allows feature selection and adjustment of coefficients for unnecessary features using penalties like L1, L2, or Elastic-Net.

K-nearest Neighbors (KNN) is a simple, ^15^ instance-based learning model that computes the distance between a new observation and existing instances, using the ‘K’ closest instances for prediction. It’s adaptable to input changes and works well with multi-class cases and irregular decision boundaries.

Support Vector Machine (SVM) is effective in high-dimensional spaces, ^16^ seeking a hyperplane with the maximum margin for classification. It uses the kernel trick for non-linear decision boundaries and is robust against overfitting but can be computationally intensive.

Random Forests, ^17^ an ensemble model, constructs many decision trees and outputs the mode or mean prediction of these trees. It introduces randomness in training and considers a random subset of features for splitting, making it effective against overfitting and capable of handling many input variables.

XGBoost, ^18^ based on gradient boosting, uses decision tree ensembles and systematically corrects errors, offering fast computation and a regularization parameter to minimize overfitting. It’s adaptable to various prediction tasks with custom optimization objectives.

LightGBM grows trees leaf-wise and employs Gradient-based One-Side Sampling (GOSS) and Exclusive Feature Bundling (EFB) for efficiency and scalability.^19^ It’s more accurate but can overfit on smaller datasets.

Lastly, 1D-CNN, adapted from Convolutional Neural Networks for sequential data like time series and natural language. ^20^ It captures local patterns in sequences, handles long sequences efficiently, and extracts local features effectively.

### 2.3. Hyperparameter Optimization

Hyperparameter optimization is a crucial step that impacts the success of a model in machine learning. Hyperparameters are set before the training process and significantly influence a model’s learning effectiveness. The optimization process involves searching for the best hyperparameter configurations, which can be computationally intensive and time-consuming.

This study used Hyperopt as an efficient tool for streamlining hyperparameter optimization. ^11^ Hyperopt uses the Tree-structured Parzen Estimator (TPE) method, which constructs a probabilistic model of the objective function to guide the search, leading to a more efficient process and often superior results. Hyperopt can be easily integrated into machine learning workflows, significantly reducing the resources and time required for optimization and improving the model’s overall effectiveness. Table A in the Appendix shows the hyperparameter searching spaces of each model.

### 2.4. Machine Learning-based Ensemble Model (Ensemble (ML))

This study employed the loss function-based approach as the machine learning-based ensemble model (Ensemble (ML)). This technique optimized the combination of models by leveraging a loss function, which measured the bias of predicted values from actual values.

In the approach, each base model in the ensemble was assigned a weight based on its performance as measured by the cross-entropy loss function. ^21^ The ensemble then combined these models’ predictions according to their weights to make the final prediction, enhancing prediction accuracy. The formulas Eq. 1 and Eq. 2 are shown below:

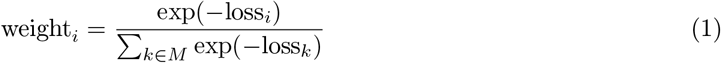

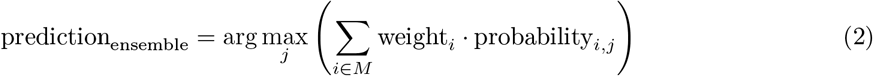

where *M* represents the set of models used in the ensemble, *i* denotes an individual model within this ensemble, weight_*i*_ is the weight assigned to model *i*, probability_*i,j*_ is the probability assigned to class *j* by model *i*, and *j* represents the class. The function arg max_*j*_ selects the class *j* for which the sum of weighted probabilities across all models is maximized.

In essence, ensemble models utilize a mechanism to fine-tune the combination of multiple models. This provided an optimized way of ensemble learning, thus improving prediction performance.

### 2.5. Clinical Risk Stratification Recommendations by the Combination of Ensemble Model and ELN 2022 (Ensemble (ML+ELN))

After predicting by the Ensemble (ML) model, the study proposed clinical risk stratification recommendations by considering the Ensemble (ML) model and ELN 2022 risk prediction (Ensemble (ML+ELN)). The proposed clinical risk stratification recommendations pipeline is shown in Fig. 2.

**Figure 2.**
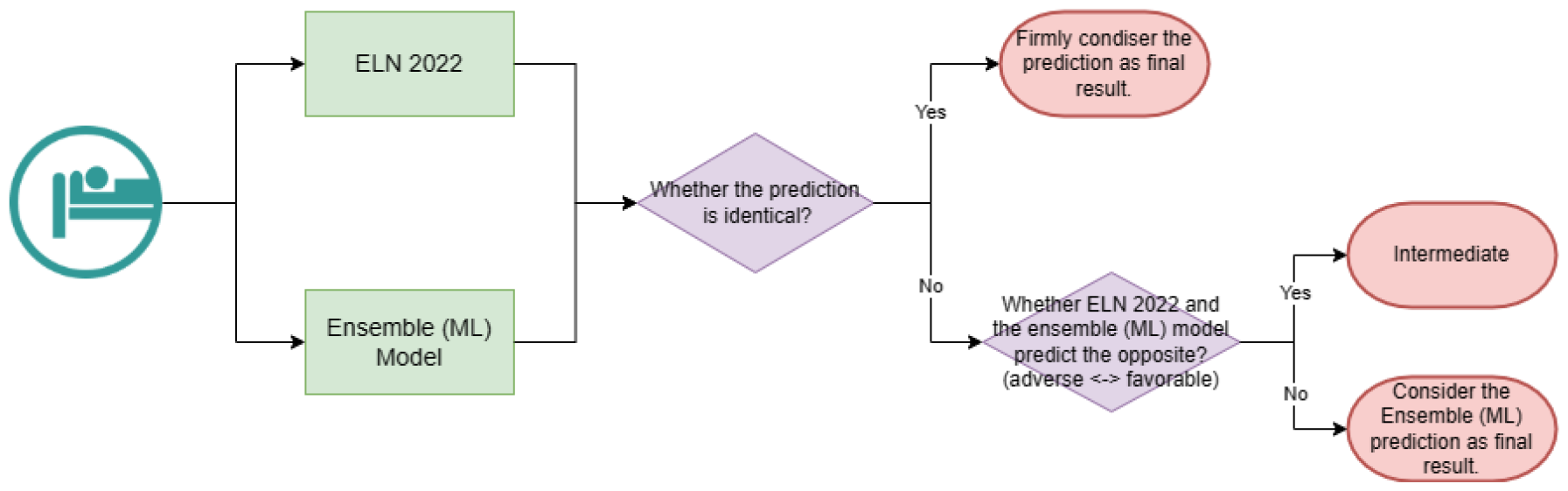
The proposed clinical risk stratification used in the Ensemble (ML+ELN) model.

The Ensemble (ML+ELN) model considered patients’ risk levels by the following step.

1. The ELN 2022 and Ensemble (ML) models predict patient risk levels, and the Ensemble (ML+ELN) model then confidently predicts those patients with the same risk level.
2. For samples contrary to the prediction, the Ensemble (ML+ELN) model considered them intermediate. That is to say; for samples predicted as adverse by ELN 2022 and favorable by the Ensemble (ML) model or those predicted by ELN 2022 as adverse and favorable by the Ensemble (ML) model, the Ensemble (ML+ELN) model treated them as intermediate.
3. The rest treated the Ensemble (ML) prediction as the final result.

## 3. Results

The study focused on enhancing risk stratification in AML patients using the proposed model, Ensemble (ML+ELN). The model was created using a dataset of 1,213 AML patients from the National Taiwan University Hospital (NTUH) and subsequently tested on the NCRI cohort to assess its generalizability. ^7;8;9;10^ In addition, to ensure the robustness of the model, it was developed by conducting 50 experiments on the NTUH dataset with different partitions of training and validation sets, and then tested on the external NCRI cohort.

### 3.1. Performance on the validation set

To ensure the model’s robustness, this study conducted 50 independent experiments, each establishing a model with a distinct partition of the training and validation sets on the NTUH cohort.

In one experiment, its validation set is used to compare the proposed Ensemble (ML+ELN) model with the ELN 2022, the Ensemble (ML), and seven machine learning models, including logistic regression, k-nearest neighbor (KNN), support vector machine (SVM), random forest, XGBoost, LightGBM, and 1D-CNN. It is shown in Table 2 that Ensemble (ML+ELN) outperformed the other models on the weighted F1 score and the concordance index (c-index). ^22^ The ML+ELN model is also superior in distinguishing the adverse risk level from the intermediate (p-value for logrank test) (Fig. 3). ^22;23^ In Table 3, we further examined a subset of patients with identical predicted risk levels from ELN and the Ensemble (ML) models. All the metrics are largely improved when compared to Table 2. Finally, the model with the best c-index among the 50 experiments is released for future prediction (Data Sharing Statement). Different presentations of the released model on the validation set, including survival curves, confusion matrices, and distribution flows of samples, are shown in Fig. 4. The p-value for logrank test of the adverse curve against the intermediate curve is much smaller in Ensemble (ML+ELN). The confusion matrices, as well as the distribution flows of samples, also demonstrated that the ML+ELN model infrequently misclassified the favorable or intermediate samples as adverse.

**Table 2.** The performance of the 50 models on the corresponding validation set (n=161) with a mean and standard deviation in 50 times experiments.

| Model | Accuracy | Weighted F1 score<sup>1</sup> | c-index<sup>22</sup> |
| --- | --- | --- | --- |
| **ELN 2022** | 0.49 ± 0.03 | 0.49 ± 0.03 | 0.63 ± 0.02 |
| **Logistic Regression** | 0.48 ± 0.03 | 0.47 ± 0.03 | 0.64 ± 0.02 |
| **KNN** | 0.45 ± 0.03 | 0.45 ± 0.03 | 0.62 ± 0.02 |
| **SVM** | 0.48 ± 0.03 | 0.48 ± 0.03 | 0.64 ± 0.02 |
| **Random Forest** | 0.49 ± 0.03 | 0.48 ± 0.03 | 0.63 ± 0.02 |
| **XGBoost** | 0.47 ± 0.03 | 0.47 ± 0.03 | 0.62 ± 0.02 |
| **LightGBM** | 0.48 ± 0.03 | 0.48 ± 0.03 | 0.63 ± 0.02 |
| **1D-CNN** | 0.46 ± 0.04 | 0.45 ± 0.04 | 0.62 ± 0.02 |
| **Ensemble (ML)** | **0.50 ± 0.03** | 0.49 ± 0.03 | 0.64 ± 0.02 |
| **Ensemble (ML + ELN)** | 0.49 ± 0.03 | **0.50 ± 0.03** | **0.65 ± 0.02** |

<sup>1</sup> The weighted F1 score calculates the F1 score for each class independently and uses a weight that depends on the number of true instances for each class to average the prediction.

**Table 3.**
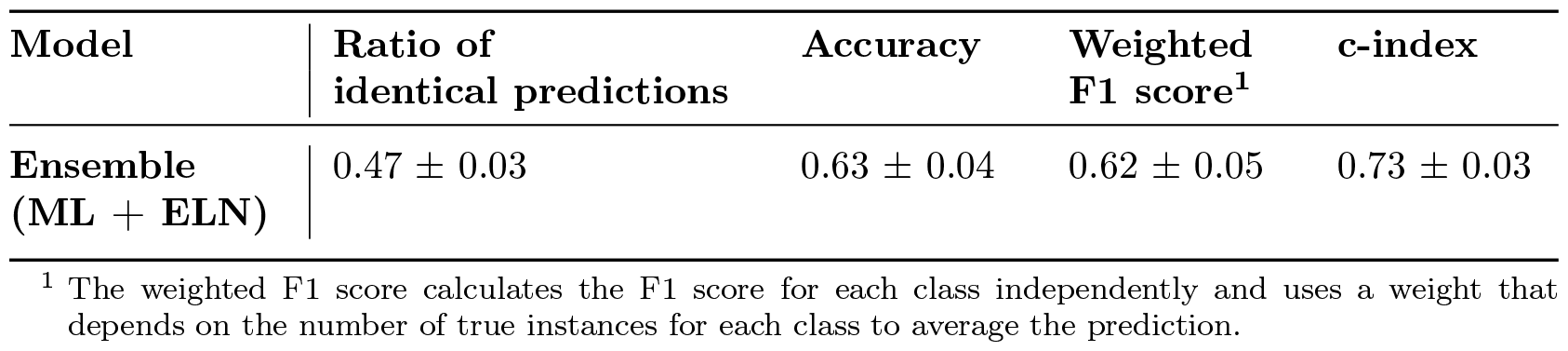
The performance of the Ensemble (ML+ELN) model for the samples where the Ensemble (ML) model and ELN 2022 predict identical risk levels. The values are mean and standard deviation in 50 times of experiments.

| Model | Ratio of identical predictions | Accuracy | Weighted F1 score<sup>1</sup> | c-index |
| --- | --- | --- | --- | --- |
| Ensemble (ML + ELN) | $0.47 \pm 0.03$ | $0.63 \pm 0.04$ | $0.62 \pm 0.05$ | $0.73 \pm 0.03$ |

<sup>1</sup> The weighted F1 score calculates the F1 score for each class independently and uses a weight that depends on the number of true instances for each class to average the prediction.

**Figure 3.**
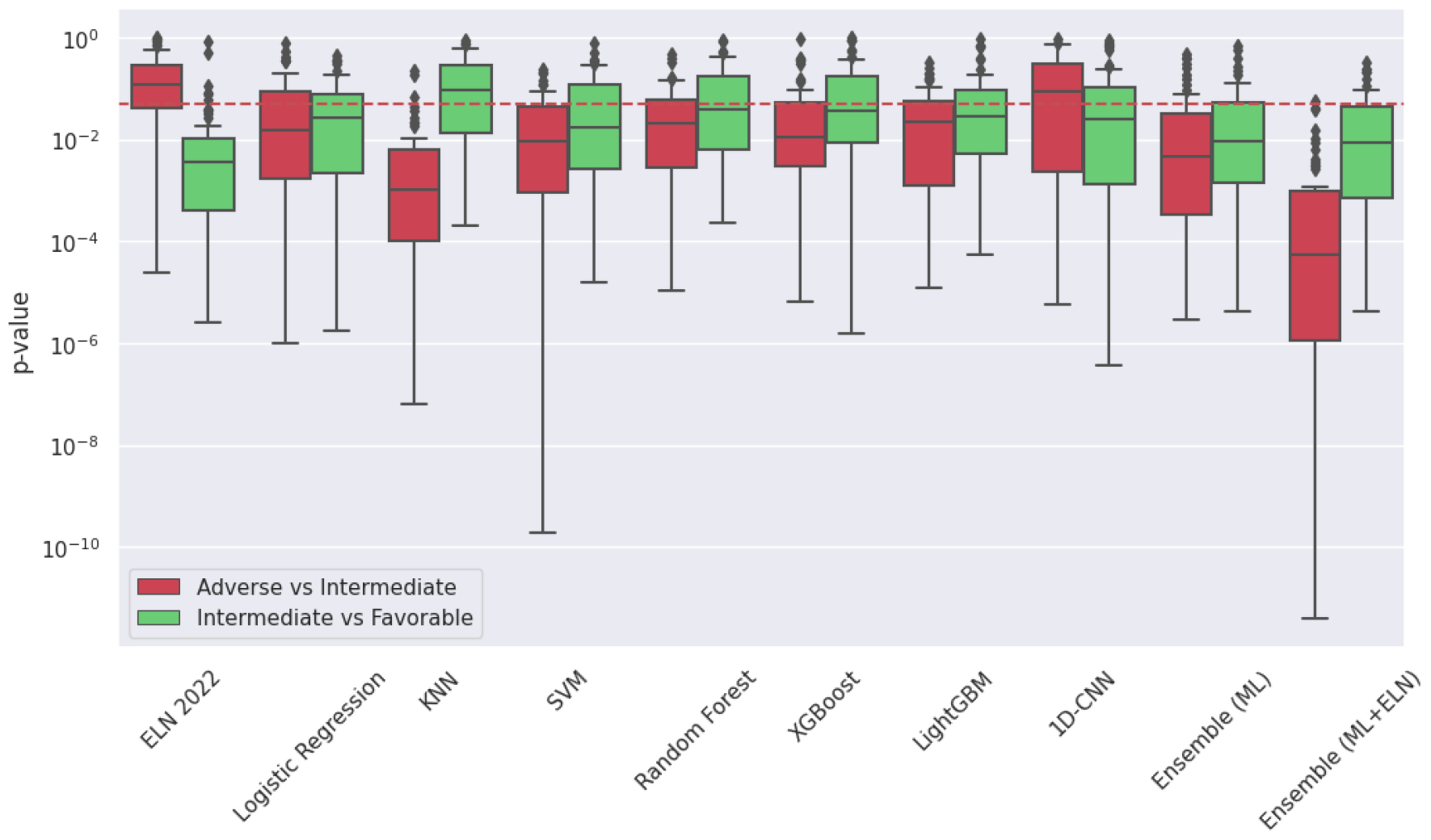
Distribution of p-values for logrank test between different survival curves for 50 experiments in different models on the validation set. The red dashed line has a p-value of 0.05.

**Figure 4.**
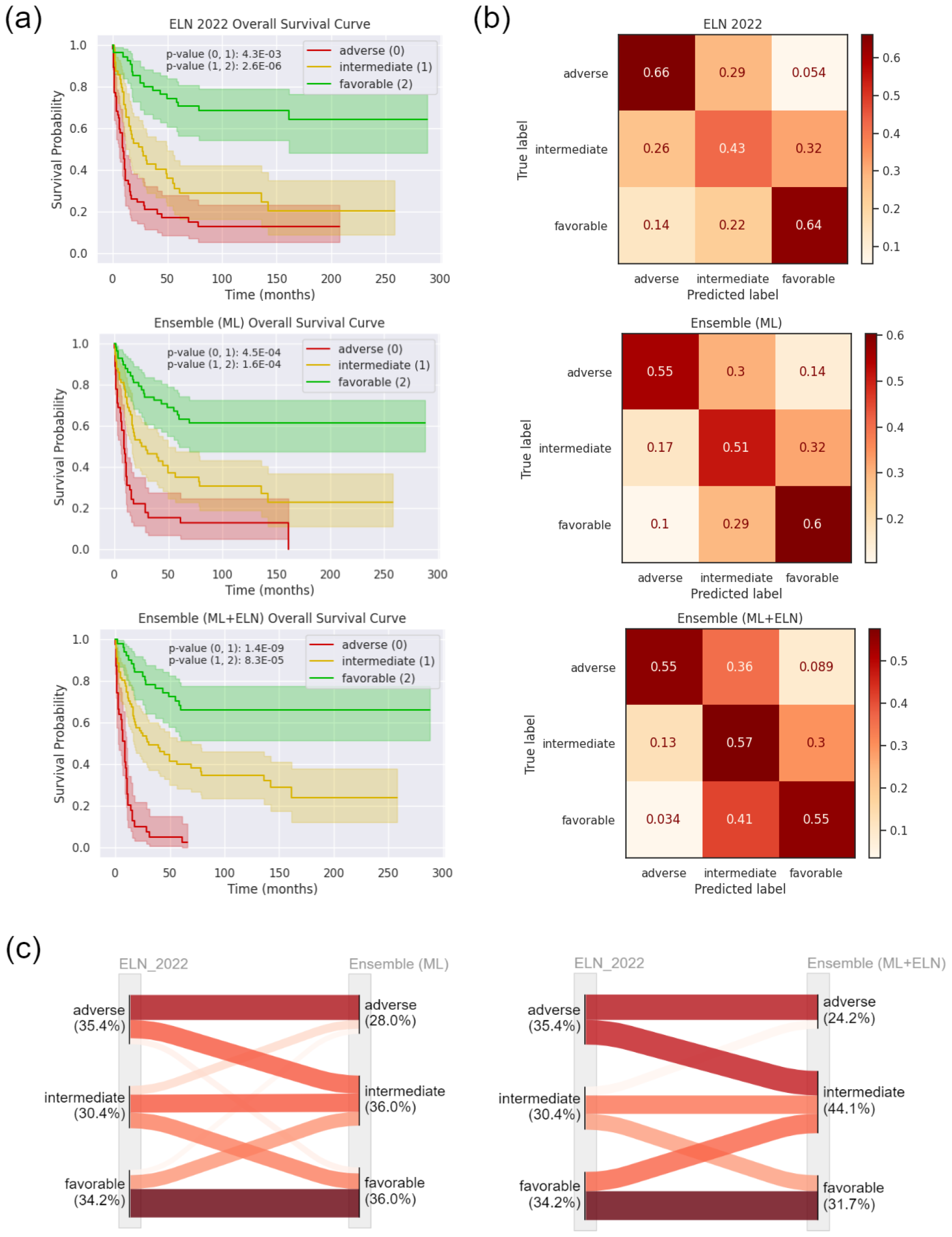
Different presentations of the released model on the validation set. (a) Survival curves. (b) Confusion matrices. (c) Distribution flows of samples.

### 3.2. Performance on the external cohort

Next, we tested the 50 models built from NTUH on the NCRI cohort for performance evaluation. The Ensemble (ML+ELN) model performed the best on all evaluation metrics on the external cohort, as described as follows. The proposed model showcased its effectiveness in improving an average concordance index (c-index) from 0.61 (ELN) to 0.64 (ML+ELN) (Table 4), ^22^ and the median hazard ratios: ^24^ 1.85 (ELN) to 2.2 (ML+ELN) for adverse versus intermediate groups (18% improved), 1.53 (ELN) to 1.97 (ML+ELN) for intermediate versus favorable groups (29% improved), and 2.83 (ELN) to 4.24 (ML+ELN) for adverse versus favorable groups (50% improved) (Fig. 5a). Furthermore, it demonstrated the capability to differentiate between pairs of risk levels. The distribution of 50 p-values are plotted in Fig. 5b. The median improves from 1.37e-22 (ELN) to 3.01e-41 (ML+ELN) for adverse versus intermediate groups and 8.31e-8 to 2.35e-19 for intermediate versus favorable groups. Besides, different presentations of the released model on the external cohort are shown in Fig. 6. Compared to the performance of released model in validation set, the ability to differentiate intermediate and favorable

**Table 4.** The performance on the NCRI cohort.

| Model | c-index with mean and standard deviation in 50 experiments<sup>1</sup> | c-index for the released models <sup>2</sup> |
| --- | --- | --- |
| ELN 2022 | 0.61 ± 0.00 | 0.612 |
| Ensemble (ML) | 0.64 ± 0.01 | 0.642 |
| Ensemble (ML+ELN) | **0.64 ± 0.00** | **0.645** |

<sup>1</sup> Each model was trained in 50 experiments on the NTUH cohort, using different training and validation partitions. The whole NCRI cohort was used to test the performance in each experiment.
<sup>2</sup> Released models were selected through the best c-index on the validation set in 50 experiments.

**Figure 5.**
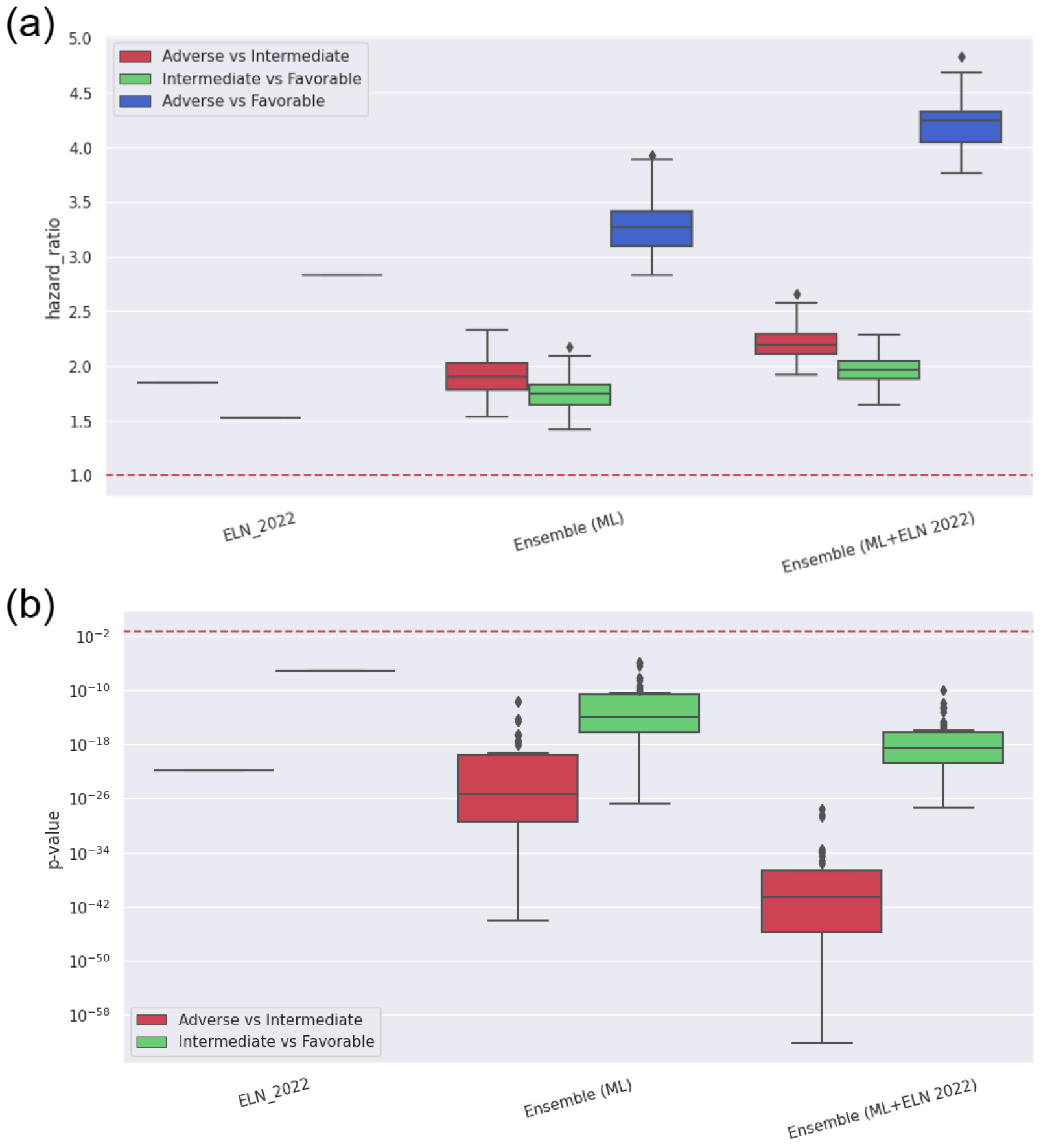
Effect sizes in different models on the external cohort. (a) Distribution of hazard ratios for different risk levels for 50 experiments in different models. The red dashed line has a hazard ratio of 1. (b) Distribution of p-values for logrank test between different survival curves for 50 experiments in different models on the external cohort. The red dashed line has a p-value of 0.05.

**Figure 6.**
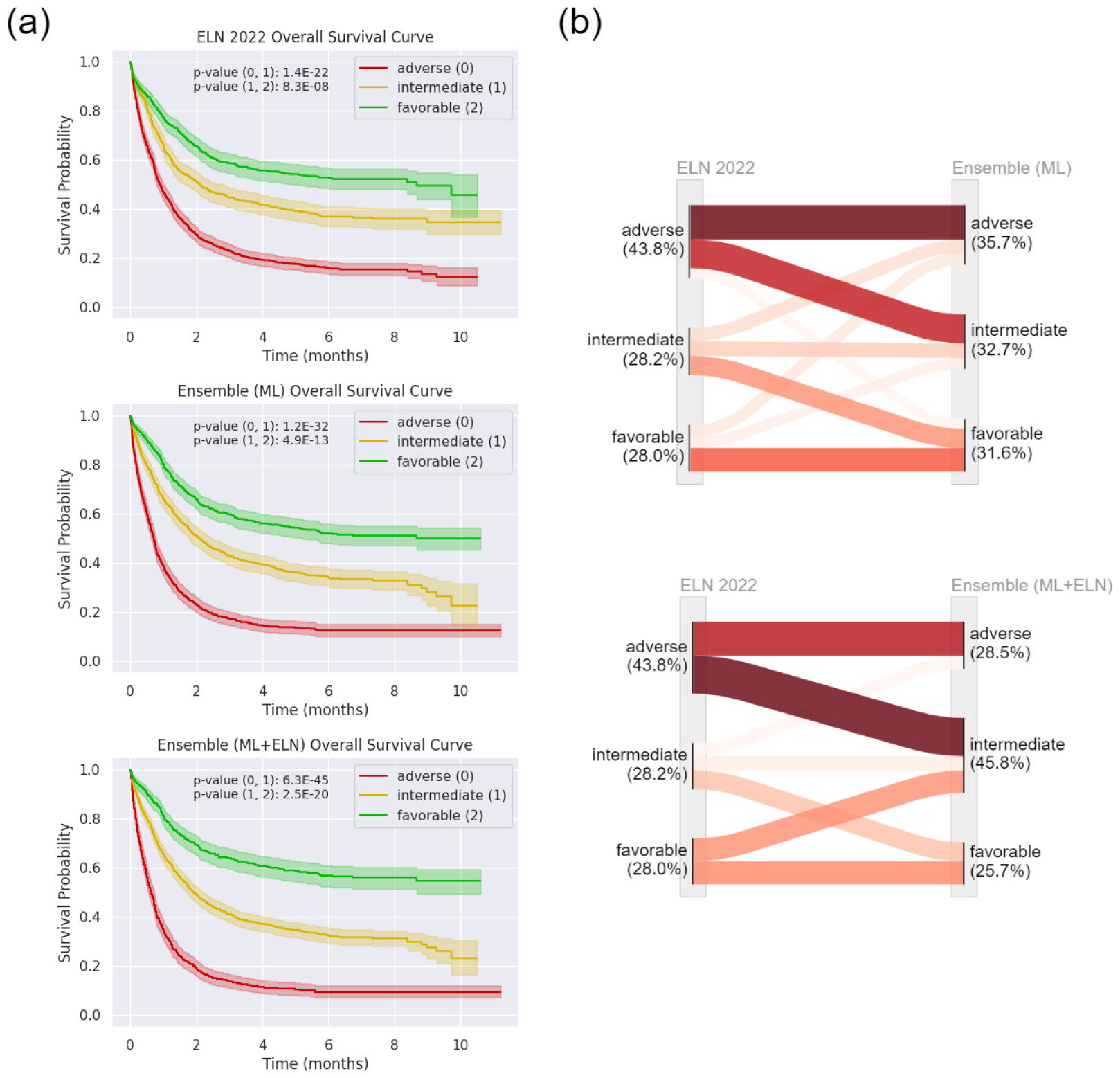
Different presentations of the released model on the NCRI cohort. (a) Survival curves. (b) Distribution flows of samples.

### 3.3. Insights for patients predicted as adverse by ELN 2022

Here, we partitioned the patients predicted as adverse by ELN into two groups. One group included samples classified as adverse by both the ELN 2022 and ensemble (ML+ELN) model, while the other consisted of samples identified as adverse by ELN 2022 but not by the ensemble (ML+ELN) model. The survival curves for these groups are illustrated in Fig. 7a. It is observed that the survival curves of these two groups are far apart. Distribution of p-values for logrank test between the survival curves of the two groups suggested that the Ensemble (ML+ELN) model can further identify patients with higher risk (Fig. 7b). ^22;23^

**Figure 7.**
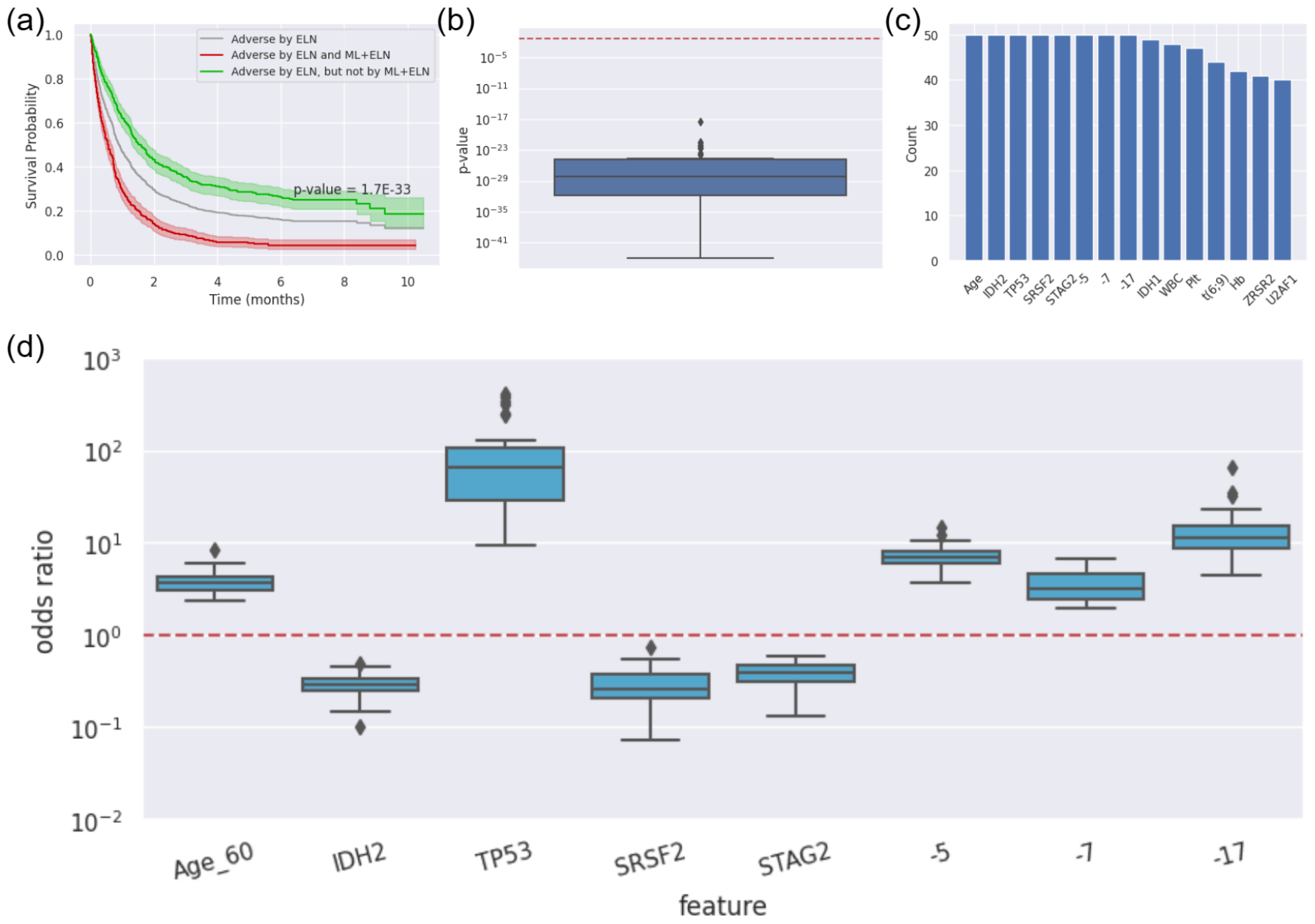
Biomarkers for further distinguishing more favorable patients from the NCRI patients identified as adverse according to the ELN 2022 guidelines. (a) Survival curves for the released models where patients were classified as adverse by ELN 2022. The p-value results from the logrank test between the red and green survival curves. (b) Distribution of p-values for logrank test between the survival curves of two patient groups in 50 experiments, where one group is classified as adverse by both ELN 2022 and an Ensemble (ELN+ML) model, and the other is classified as adverse by ELN 2022 but non-adverse by the Ensemble (ELN+ML) model. The red dashed line has a p-value of 0.05. (c) The Significant biomarkers (p-value<0.05) were observed over 40 of 50 experiments. (d) Analyzing the effect of consistently occurred significant biomarkers by an odds ratio. ^27^ The red dashed line has an odds ratio of 1.

We further identified significantly differential features (p-value < 0.05) in between two groups using different statistical tests: T-tests for numerical features and chi-square tests for categorical ones. ^25;26^ For the external cohort, eight biomarkers consistently appeared in all 50 experiments as the significant features (Fig. 7c). An odds ratio was used to demonstrate the impact of these significant features on risk prediction (Fig. 7d). ^27^ The feature ‘age’ was categorized with a threshold of 60 years (Age_60). Features like Age_60, *TP53*, and specific karyotypes (-5, -7, -17) led the Ensemble (ML+ELN) model to categorize patients as adverse. In contrast, features such as *IDH2, SRSF2*, and *STAG2* led to a non-adverse prediction. In other words, for patients identified as adverse in ELN 2022, determining if they are older than 60, carrying *TP53*, or possessing the aforementioned karyotypes can help further identify patients with a higher adverse risk. On the other hand, checking for *IDH2, SRSF2*, or *STAG2* can help identify patients with a less adverse risk.

### 3.4. Insights for patients predicted as favorable by ELN 2022

Similarly, we partitioned the patients predicted as favorable by ELN into two groups. One group included samples classified as favorable by both the ELN 2022 and ensemble (ML+ELN) model, while the other consisted of samples identified as favorable by ELN 2022 but not by the ensemble (ML+ELN) model. The survival curves of one particular experiment for these groups are illustrated in Fig. 8a. It is observed that the survival curves of these two groups are far apart. Distribution of the p-values for logrank test between the survival curves of the two groups in 50 experiments suggested that the Ensemble (ML+ELN) model can further identify patients with low risk (Fig. 8b). ^22;23^

**Figure 8.**
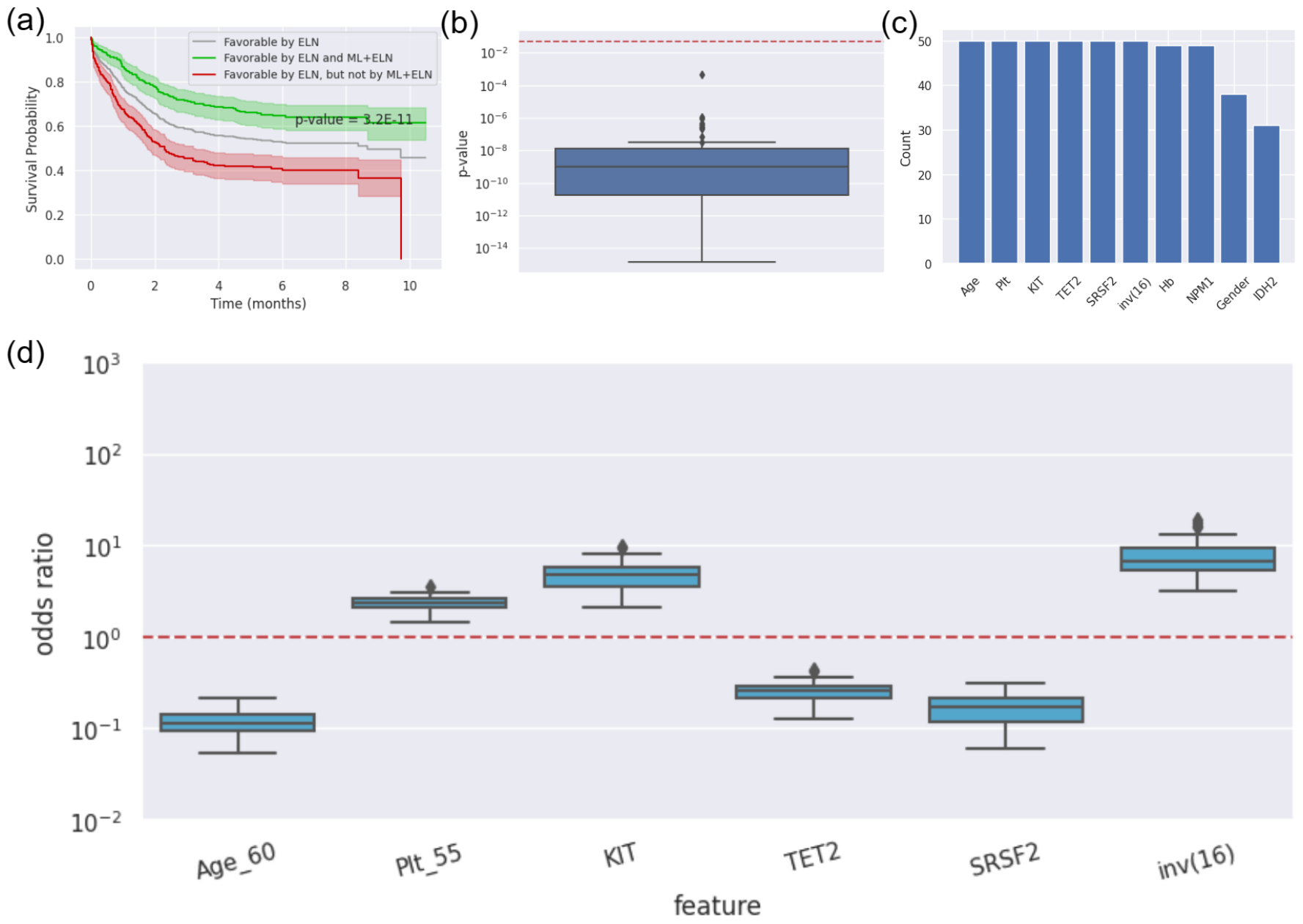
Biomarkers for further distinguishing more favorable patients from the NCRI patients identified as favorable according to the ELN 2022 guidelines. (a) Survival curves for the released models where patients were classified as favorable by ELN 2022. The p-value is the logrank test between the red and green survival curves. (b) Distribution of p-values for logrank test between the survival curves of two patient groups in 50 experiments, where one group is classified as favorable by both ELN 2022 and an Ensemble (ELN+ML) model, and the other is classified as favorable by ELN 2022 but non-favorable by the Ensemble (ELN+ML) model. The red dashed line has a p-value of 0.05. (c) The Significant biomarkers (p-value<0.05) were observed over 30 of 50 experiments. (d) Analyzing the effect of consistently occurred significant biomarkers by an odds ratio. ^27^ The red dashed line has an odds ratio of 1.

Afterward, we assessed significantly differential features (p-value < 0.05) in two groups using the same statistical tests described in section 4.3. For the external cohort, six biomarkers consistently appeared in the 50 experiments as the significant features (Fig. 8c). In the odds ratio metric (Fig. 8d), ^27^ the feature ‘age’ was categorized with a threshold of 60 years (Age_60), and ‘Plt’ (Platelets) was categorized with a threshold of 55 thousand per microliter (Plt_55). Features like Plt_55, *KIT*, and inv(16) led the Ensemble (ML+ELN) model to categorize patients as favorable. In contrast, features such as Age_60, *TET2*, and *SRSF2* led to a non-favorable prediction. Therefore, for patients identified as favorable in ELN 2022, determining if they are younger than 60, the concentration of platelets is more than 55 thousand per microliter, or patients carry *KIT* or inv(16) can help further identify patients with a more favorable risk. On the other hand, checking for *TET2* or *SRSF2* can help identify a less favorable group.

### 3.5. Variations in Hematopoietic Stem Cell Transplantation Effectiveness Across Different Risk Levels

The study assessed the impact of hematopoietic stem cell transplantation (HSCT) on patients classified into different risk levels as part of the NCRI cohort. Of the 2,113 patients in the cohort, 527 underwent HSCT. The study utilized hazard ratios to analyze outcomes based on risk assessments made by two different models: the ELN 2022 criteria and the Ensemble (ML) model (Fig. 9a). Patients described in Fig. 9c and Fig. 9d are recommended for the HSCT treatment.

**Figure 9.**
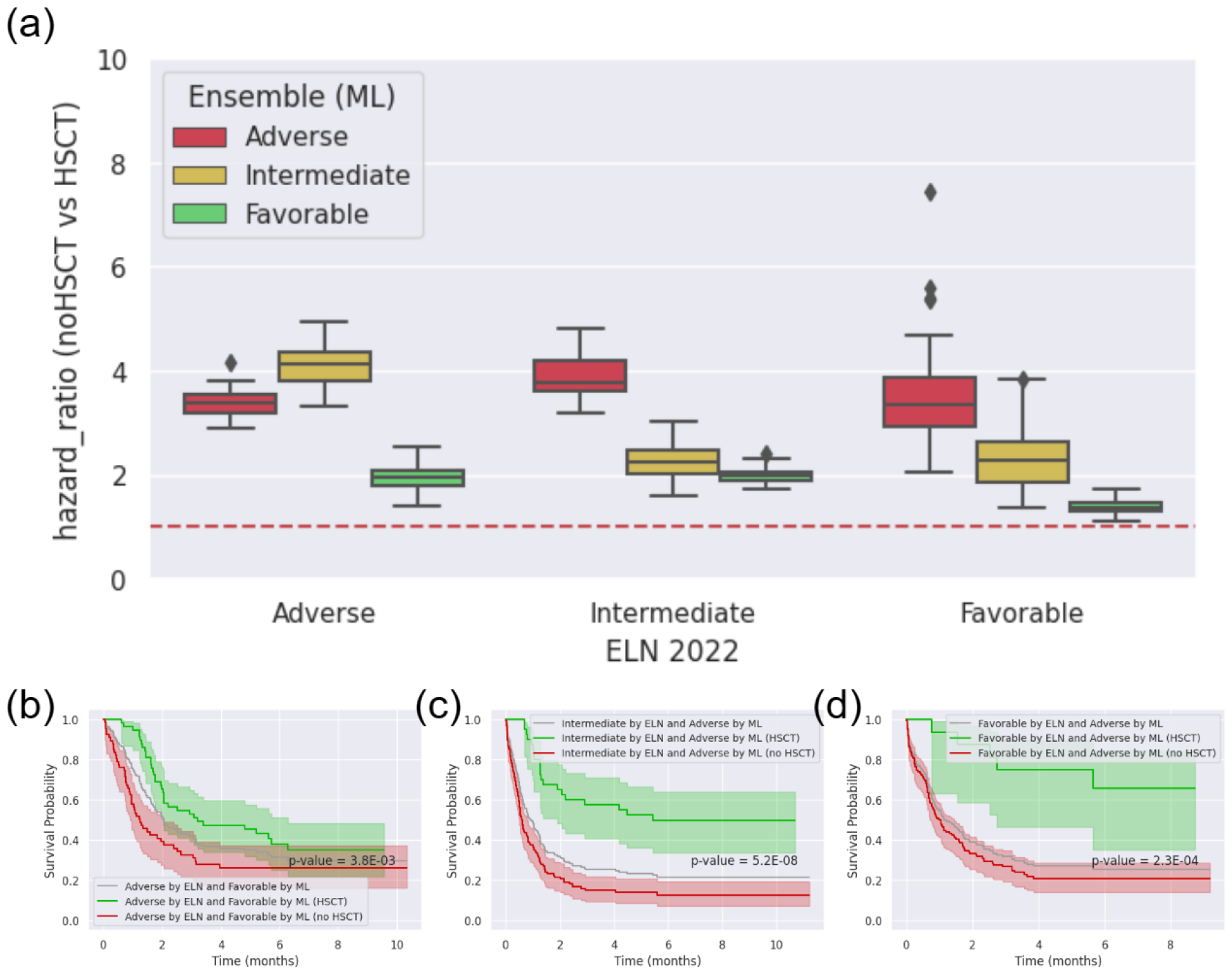
Comparative analysis of the effectiveness of HSCT according to ELN 2022 and the Ensemble (ML) model in predicting different risk levels on the external cohort. (a) Overall HSCT effectiveness on the NCRI cohort. The red dashed line has a hazard ratio of 1. (b) Survival curves for patients predicted as adverse by ELN 2022 but favorable by the released Ensemble (ML) model. The p-value is the logrank test between the red and green survival curves. (c) Survival curves for patients predicted as intermediate by ELN 2022 and adverse by the released Ensemble (ML) model. The p-value is the logrank test between the red and green survival curves. (d) Survival curves for patients predicted as favorable by ELN 2022 but adverse by the released Ensemble (ML) model. The p-value is the logrank test between the red and green survival curves.

The findings revealed notable variations in the effectiveness of HSCT depending on the risk level predictions made by these two models. Specifically:

1. For patients categorized as adverse according to ELN 2022 but favorable by the Ensemble (ML) model, HSCT showed a reduced benefit. This HSCT effectiveness was comparable to, or even less than, that observed in patients deemed favorable by ELN 2022. The survival curve from the released models for these patients is shown in Fig. 9b.
2. Patients rated as intermediate-risk by ELN 2022 and adverse by the Ensemble (ML) model showed a level of HSCT effectiveness similar to those patients categorized as adverse by ELN 2022. The survival curve from the released models for these patients is shown in Fig. 9c.
3. Patients identified as favorable by ELN 2022 but adverse by the Ensemble (ML) model experienced a significantly enhanced benefit from HSCT. This effectiveness was similar to or greater than that seen in patients classified as adverse by ELN 2022. The survival curve from the released models for these patients is shown in Fig. 9d.

Our study expands on prior research in HSCT outcomes by further dividing risk level groups, incorporating both ELN 2022 criteria and Ensemble (ML) model predictions. This approach offers a more detailed risk assessment, particularly identifying a subset that responds variably to HSCT.

## 4. Discussion

This study presents a significant advancement in the risk stratification of Acute Myeloid Leukemia (AML) patients through an ensemble machine learning model. This model, integrated with the European LeukemiaNet (ELN) 2022 recommendations, has shown remarkable effect in accurately categorizing AML patients into distinct risk groups.

The study revealed key genetic and clinical features influencing AML prognosis through statistical analysis. Integrating the findings from the previous literature with the study offers a comprehensive understanding of risk stratification in Acute Myeloid Leukemia (AML) patients. From our insights for patients predicted as adverse by ELN, we discovered significant biomarkers, including age more than 60, *TP53* mutation, and karyotypes (-5, -7, -17) linked with higher risk and mutations such as *IDH2, SRSF2*, and *STAG2* indicated a lower risk. Previous studies have shown that older AML patients exhibit distinct genetic alterations compared to younger patients, with mutations like *TP53* being poor prognostic factors. ^28;29^ In addition, the *TP53* mutation, known for its stability during AML evolution, is associated with poor prognosis, especially in patients with complex karyotypes. ^29^ Contrarily, mutations in the cohesin complex gene *STAG2* have contributed to the disease’s complexity and showed a trend toward improved long-term outcomes. ^30^ Moreover, the earlier research highlighted the prognostic significance of various genetic alterations and cytogenetic profiles, such as the favorable impact of *IDH2* mutations and the negative implications of an unfavorable karyotype (-5, -7, -17) on treatment outcomes and survival rates in AML. ^31^

Similarly, from our insights for patients predicted as favorable by ELN, biomarkers like a platelet count above 55,000 per microliter, *KIT* mutations, and inv(16) karyotype were associated with favorable outcomes, whereas age more than 60 and mutations such as *TET2* and *SRSF2* suggested a less favorable prognosis. A previous study has shown that older AML patients exhibit distinct genetic profiles with mutations like *DNMT3A* and *TP53* being poor prognostic indicators. ^28^ Platelet count at diagnosis is also a critical factor influencing treatment outcomes. ^32^ The findings regarding *TET2* mutation in acute myeloid leukemia (AML) patients, particularly those with intermediate-risk cytogenetics, underscore its role as an unfavorable prognostic factor. ^33^ Moreover, myelodysplasia-related mutations like *SRSF2* indicate poor outcomes across all age groups, underscoring the need for intensive treatment. ^34^ Another earlier research highlighted the prognostic importance of favorable karyotypes inv(16) in AML. ^31^

The analysis of HSCT effectiveness across different risk levels offers novel perspectives on treatment effects. The study’s findings highlight the model’s potential to identify subsets of patients who might benefit more or less from HSCT, thus offering treatment recommendations and improving outcomes. While these findings are significant, they should be interpreted cautiously due to potential selection biases in retrospective studies. Prospective studies are needed to validate and further explore these insights.

In conclusion, the application of this model promises to enhance existing prognostic assessments and aid in personalized treatment planning. Integrating advanced machine learning techniques with established medical guidelines opens new avenues for enhancing patient care in AML and potentially other complex diseases.

## Data Availability

The data generated during and/or analyzed during the current study are available from the corresponding author upon reasonable request.
The UK-NCRI dataset in the present study is available online at https://www.nature.com/articles/s41467-022-32103-8#Sec1.
The Ensemble (ML+ELN) and Ensemble (ML) models are available at https://github.com/hardness1020/AML_Risk_Stratification.

https://ega-archive.org/studies/EGAS00001000570

https://www.nature.com/articles/s41467-022-32103-8#Sec1

https://github.com/hardness1020/AML_Risk_Stratification

## 5. Contributors

MSC conceived the main studies. MSC, CHT, and CYC contributed to the conception and design of the main studies. MSC, CHT, WCC, HFT, and HAH contributed to data collection. CHT, WCC, HFT, and HAH offered medical insights. MSC wrote the draft of the manuscript. CYC revised and edited the manuscript. All authors approved the final version and had full access to all the data.

## 6. Declaration of interests

No conflicts of interest exist.

## 7. Data Sharing Statement

This retrospective study received approval from the NTUH Research Ethics Committee. Written informed consent was obtained from all participants in accordance with the Declaration of Helsinki, under the approval number 201709072RINC. The Ensemble (ML+ELN) and Ensemble (ML) models are available in: https://github.com/hardness1020/AML_Risk_Stratification

---

**Table A.** The hyperparameters searching space of each model.

| Model | Hyperparameter Searching Space |
| --- | --- |
| Logistic Regression | 'C': hp.loguniform('C', -5, 0),<br>'penalty': hp.choice('penalty', ['l1', 'l2']),<br>'solver': hp.choice('solver', ['liblinear']),<br>'tol': hp.loguniform('tol', -4, -2),<br>'max_iter': hp.choice('max_iter', [1000]),<br>'random_state': hp.choice('random_state', [0]) |
| KNN | 'n_neighbors': hp.choice('n_neighbors', [5, 10, 15, 20, 25, 30]),<br>'weights': hp.choice('weights', ['uniform', 'distance']),<br>'algorithm': hp.choice('algorithm', ['auto', 'ball_tree']),<br>'leaf_size': hp.choice('leaf_size', [20, 30, 40]),<br>'p': hp.choice('p', [1, 2]) |
| Support Vector Machine | 'C': hp.loguniform('C', -5, 2),<br>'kernel': hp.choice('kernel', ['poly', 'rbf', 'sigmoid']),<br>'gamma': hp.choice('gamma', ['scale', 'auto']),<br>'coef0': hp.uniform('coef0', 0, 1),<br>'tol': hp.loguniform('tol', -4, -2),<br>'cache_size': hp.choice('cache_size', [2000]),<br>'shrinking': hp.choice('shrinking', [True, False]),<br>'break_ties': hp.choice('break_ties', [False, True]),<br>'class_weight': hp.choice('class_weight', [None, 'balanced']),<br>'probability': hp.choice('probability', [True]) |
| Random Forest | 'n_estimators': hp.choice('n_estimators', np.arange(100, 1001, 100, dtype=int)),<br>'criterion': hp.choice('criterion', ['gini']),<br>'max_depth': hp.choice('max_depth', np.arange(5, 20, dtype=int)),<br>'min_samples_split': hp.choice('min_samples_split', np.arange(2, 11, dtype=int)),<br>'min_samples_leaf': hp.choice('min_samples_leaf', np.arange(1, 11, dtype=int)),<br>'min_weight_fraction_leaf': hp.uniform('min_weight_fraction_leaf', 0, 0.5),<br>'min_impurity_decrease': hp.uniform('min_impurity_decrease', 0, 0.5),<br>'class_weight': hp.choice('class_weight', [None, 'balanced']),<br>'n_jobs': hp.choice('n_jobs', [32]),<br>'max_features': hp.choice('max_features', ['sqrt']) |
| XGBoost | 'eta': hp.loguniform('eta', -7, 0),<br>'max_depth': hp.choice('max_depth', np.arange(1, 11, dtype=int)),<br>'subsample': hp.uniform('subsample', 0.2, 1),<br>'colsample_bytree': hp.uniform('colsample_bytree', 0.2, 1),<br>'colsample_bylevel': hp.uniform('colsample_bylevel', 0.2, 1),<br>'min_child_weight': hp.loguniform('min_child_weight', -16, 2),<br>'alpha': hp.uniform('alpha', 0, 1),<br>'lambda': hp.uniform('lambda', 0, 1),<br>'gamma': hp.uniform('gamma', 0, 1) |
| LightGBM | 'learning_rate': hp.loguniform('learning_rate', -5, -2),<br>'max_depth': hp.choice('max_depth', np.arange(3, 11, dtype=int)),<br>'num_leaves': hp.choice('num_leaves', np.arange(8, 51, dtype=int)),<br>'min_data_in_leaf': hp.choice('min_data_in_leaf', np.arange(10, 20, dtype=int)),<br>'verbose': hp.choice('verbose', [-1]) |
| 1D-CNN | 'hidden_size': hp.choice('hidden_size', [32, 64, 128]),<br>'optimizer': hp.choice('optimizer', [torch.optim.Adam, torch.optim.AdamW]),<br>'learning_rate': hp.loguniform('learning_rate', -5, -2),<br>'batch_size': hp.choice('batch_size', [32, 64, 128, 256, 512]) |

## Notes

### Competing Interest Statement

The authors have declared no competing interest.

### Funding Statement

This study did not receive any funding.

### Author Declarations

Research ethics committee of National Taiwan University Hospital gave ethical approval for this work. Written informed consent was obtained from all participants in accordance with the Declaration of Helsinki, under the approval number 201709072RINC.

### Summary of Updates

Change the order of methods and Results. Revise some sentences, making them more concise and easy to understand. Correct some errors in the figures. Add some sections.

